# Symptom onset and cellular pathology in facioscapulohumeral muscular dystrophy is accelerated by cigarette smoking

**DOI:** 10.1101/2023.05.17.23290091

**Authors:** Christopher R. S. Banerji, Philipp Heher, John Hogan, Natalie Katz, Husain Bin Haidar, Michael D. Keegan, Colin Cernik, Rabi Tawil, Ketan Patel, Peter S. Zammit, Jeffery M. Statland

**Affiliations:** The Alan Turing Institute, The British Library, 96 Euston Road, London NW1 2DB, UK; King’s College London, Randall Centre for Cell and Molecular Biophysics, New Hunt’s House, Guy’s Campus, London SE1 1UL, UK; University of Kansas Medical Center, Department of Neurology, Kansas City, KS 66160, USA; School of Biological Sciences, University of Reading, Reading, RG6 6UB, UK; Kuwait Cancer Control Centre, Ministry of Health, Kuwait; King’s College London, School of Immunology and Microbiology, Department of Infectious Diseases, Guy’s Campus, London SE1 1UL, UK; University of Rochester Medical Center, Department of Neurology, Neuromuscular Disease Unit, Rochester, New York, USA

**Keywords:** Facioscapulohumeral muscular dystrophy, FSHD, smoking, Patient registry, myoblast

## Abstract

Facioscapulohumeral muscular dystrophy (FSHD) is an incurable skeletal myopathy. In absence of therapy, lifestyle factors impacting disease progression are important for clinical management. Monozygotic twins with FSHD often exhibit dramatically different disease progression, indicating existence of environmental disease modifiers. Here we analyse the USA National Registry for Myotonic Dystrophy & Facioscapulohumeral Dystrophy, comprising 511 FSHD1 patients followed up annually for an average of 8 years. This multimodal, longitudinal dataset comprises 189 baseline and 37 annually assessed features. We developed a workflow for prospective cohort analysis and identify cigarette smoking as associated with a two-fold increase in risk of facial and lower limb involvement in FSHD1 patients. Our definition of lower limb involvement includes inability to run and climb steps unaided, important functional outcomes for FSHD patients. We then employed an assay to test the effects of cigarette smoke extract on human myoblasts in vitro. Cigarette smoke extract drove disproportionate defects in proliferation and myogenic differentiation of FSHD1 patient-derived myoblasts, compared to matched controls. Mitochondrial function was also inordinately affected in FSHD1 myoblasts exposed to cigarette smoke extract, with increased mitochondrial membrane potential and mitochondrial radical oxygen species (mitoROS) generation. Our findings support recommending smoking cessation in clinical management of FSHD.

## INTRODUCTION

Facioscapulohumeral muscular dystrophy (FSHD) is an incurable skeletal myopathy, with a prevalence of ∼12/100,000 (1). No treatment has been approved, although several compounds are in clinical trials, including a p38α inhibitor (losmapimod, Fulcrum therapeutics, NCT05397470), a latent myostatin antibody (GYM329, Hoffmann-La Roche, NCT05548556) and an antibody- RNAi hybrid (AOC, Avidity, NCT05747924). In absence of therapy, lifestyle factors impacting disease progression are clinically important. FSHD is highly heterogeneous in clinical progression, age of onset and muscle group involvement, even within families (2–7). Indeed, monozygotic twins with FSHD often demonstrate extreme diversity in clinical presentation (7). This indicates a likely role for modifiable and environmental factors in FSHD disease progression, however none have yet been identified.

Heritability of FSHD is linked to epigenetic de-repression of the D4Z4 macrosatellite at chromosome 4q35 (5, 8), alongside a permissive 4qA haplotype. Epigenetic de-repression is achieved by two genetic mechanisms. In FSHD1 (OMIM: 158900, ∼95% of cases) the D4Z4 macrosatellite is truncated to 1-10 units (9), while in FSHD2 (OMIM: 158901, ∼5% of cases) patients have a mutation in an epigenetic modifier, commonly *SMCHD1* (10) but rarely *DNMT3B* (11) or *LRIF1* (12). Each 3.3kb D4Z4 unit encodes the double homeobox transcription factor DUX4. Epigenetic de-repression permits aberrant transcription of DUX4 from the distal-most D4Z4 unit, which is stabilised by splicing to a poly(A) signal on permissive 4qA haplotypes, allowing translation of DUX4 (9).

Mis-expression of DUX4 protein is believed to underlie FSHD pathogenesis. How DUX4 drives pathology remains a topic of active research with many mechanisms implicated (13–17). Among these, oxidative stress sensitivity and impaired mitochondrial function, have been identified as highly consistent features of FSHD, both in patient-derived myoblasts and healthy myoblasts over-expressing DUX4 (15, 18–22).

Clinically, FSHD classically presents as a progressive skeletal myopathy, beginning in facial muscles, before descending to specific muscles in the upper and lower body (2, 14, 23).

Although clinically evident, facial weakness is often noticed late by patients as symptoms are subtle and unobtrusive. Subsequent involvement of the shoulder girdle is symptomatic as scapular winging and impaired shoulder abduction, common presenting complaints of FSHD (24). Later in the disease process, lower limb involvement can lead to foot drop and hip flexor weakness impairing ambulation (25), and ∼20% of FSHD patients eventually require use of a wheelchair (26, 27).

While the classical presentation describes ∼75% of FSHD1 patients (2, 28), age of onset is highly variable (29) and a quarter of patients display atypical features, including facial sparing (3) and lower limb predominance (30). The search for FSHD disease modifiers, has identified genetic factors, with longer D4Z4 repeat length associated with later age of onset (2, 29, 31) and facial sparing (2, 3, 6, 32). Females have also shown later onset in some cohorts (31, 33), but earlier onset in others weighted to more severe disease (2, 34, 35). Additional modifiers, including inheritance pattern and pregnancy have been investigated, but show less consistent impact on clinical features (2, 33, 36–38). Smoking and alcohol consumption were recently examined in a cross sectional study of 198 FSHD patients, but no association was found with clinical severity (39).

Modifiable risk factors for FSHD thus remain elusive. However, current studies did not evaluate large, prospectively collected longitudinal cohorts.

Here we investigate the USA National Registry for Myotonic Dystrophy & Facioscapulohumeral Dystrophy (USA FSHD Registry), a multimodal, longitudinal dataset describing symptom evolution for 511 FSHD1 patients followed up annually for an average of 8 years (26). We develop a novel workflow for prospective analysis of this complex dataset and identify cigarette smoking as associated with a two-fold increased risk of facial and lower limb involvement in FSHD1 patients over time.

We then evaluated the effect of cigarette smoke on *in vitro* models of FSHD. Cigarette smoke medium (CSM) drives disproportionate defects in proliferation, mitochondrial function and myogenic differentiation of FSHD1 patient-derived myoblasts compared to matched controls. FSHD myoblasts are also more susceptible to CSM-induced apoptosis and exhibit inordinate increases in mitochondrial radical oxygen species (mitoROS) levels, driven by concomitant hyperpolarization of the inner mitochondrial membrane.

## RESULTS

### Pre-processing the USA FSHD registry for prospective analysis

The USA FSHD registry is a complex, multimodal, temporal data set, which required pre- processing for prospective analysis (**Fig 1A**).

**Fig. 1.**
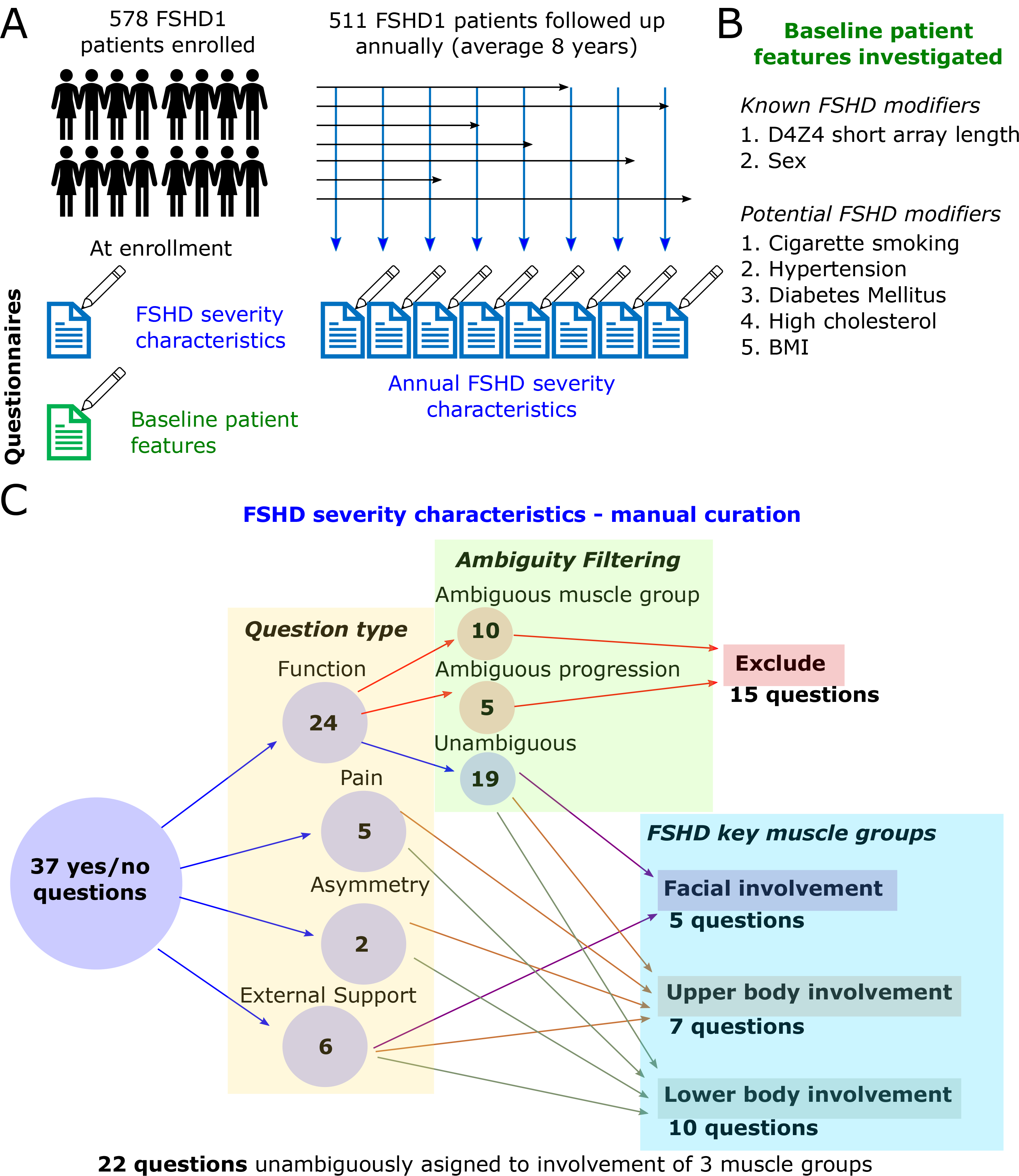
Manual curation of the USA FSHD registry. (A) Schematic showing the structure of the USA FSHD registry for prospective analysis. 578 genetically confirmed FSHD1 patients are enrolled at various ages, a baseline questionnaire completed at enrolment contains 189 features, including details of modifiable risk factors. 511/578 patients have at least 1 annual follow up, with average follow up 8 years. At each follow up a questionnaire detailing FSHD severity characteristics is completed. **(B)** Baseline patient features assessed at enrolment considered for investigation by prospective analysis. **(C)** Manual curation of 37 yes/no questions included in the annual follow up questionnaire. Questions were categorised as pertaining to muscle function, pain, asymmetry and the use of external supports, 15 questions were then excluded due to ambiguity in muscle group involvement or interpretation of the answer in relation to progression. Of the remaining 22 questions, 5 were assigned to facial involvement, 7 to upper body and 10 to lower body.

The registry comprised 578 genetically confirmed FSHD1 patients enrolled between 2001 and 2020. For each patient, 189 clinical and patient-reported features were collected by enrollment questionnaire. Features were wide ranging, spanning self-reported functional assessments, comorbidities, demographics, use of assistive devices and D4Z4 short array length, and entailed both qualitative and quantitative outputs (26).

To identify modifiable risk factors for FSHD progression present at enrollment, we focused on the five quantitative, potentially modifiable explanatory variables recorded by the enrollment questionnaire. These were BMI, self-reported diabetes, hypertension, high cholesterol and cigarette smoking, alongside the two known FSHD disease modifiers of D4Z4 short array length and sex (**Fig 1B**). Known associations between certain explanatory variables were as expected from literature (**Supplementary Table S1**).

Annual follow-up questionnaires were completed by 511 patients with an average follow up of 8 years (range: 1-17 years). We manually curated these questionnaires to extract meaningful temporal data on symptom progression for 3 key muscle groups involved in FSHD: the face, the upper body and the lower body (**Fig 1C**).

The annual questionnaire contains 24 yes/no questions covering overall physical function, 5 yes/no questions assessing distribution of muscle pain and 2 yes/no questions assessing left/right asymmetry. In addition, 4 yes/no questions cover use of ambulatory aids, one the undertaking of scapular fixation surgery and one the use of speech therapy. The questionnaire can be accessed at https://www.urmc.rochester.edu/neurology/national-registry/join.aspx.

Of these 37 yes/no questions, we attributed 5 to facial involvement, 9 to upper body involvement and 13 to lower body involvement. The remaining 10 functional questions could not be unambiguously assigned to a single muscle group and were excluded. Five functional questions attributed to upper and lower body involvement, described intermediate stages of symptom progression. Thus a ‘no’ answer could imply either an earlier or later stage of symptom, and so these 5 variables were excluded to ensure that a ‘no’ could always be interpreted unambiguously.

This resulted in 22 yes/no questions extracted from the annual follow-up questionnaire for prospective analysis: 5 describing involvement of the face, 7 upper body and 10 lower body (**Fig 1C**). For each question, reporting a ‘yes’ implied progressive involvement of the assigned muscle group.

### Workflow for prospective analysis of the USA FSHD registry

In prospective analysis with a finite population, investigation of rare events is underpowered compared to common events. To obtain a picture of common versus rare FSHD features, we ranked our 22 curated annual questions by the proportion of patients enrolled in the registry who reported ‘yes’ at any time from enrolment to latest follow up.

The 10 most common questions were affirmed by at least 60% of the cohort and span our three muscle groups of interest (**Fig 2A**). Two described facial involvement: dry eyes (affirmed by 72.1%) and trouble whistling/drinking through a straw (affirmed by 67.3%). Four described upper body involvement: asymmetrical arm weakness (affirmed by 84.7%), loss of ability to raise arms above head (affirmed by 83.4%), muscle pain in shoulder or arms (affirmed by 75.0%) and muscle pain in neck or upper back (affirmed by 68.1%). Four described lower body involvement: loss of ability to run (affirmed by 85.9%), loss of ability to climb stairs without aid of a hand rail (affirmed by 84.9%), asymmetrical leg weakness (affirmed by 78.9%) and pain in the lower back or hips (affirmed by 74.6%).

**Fig. 2.**
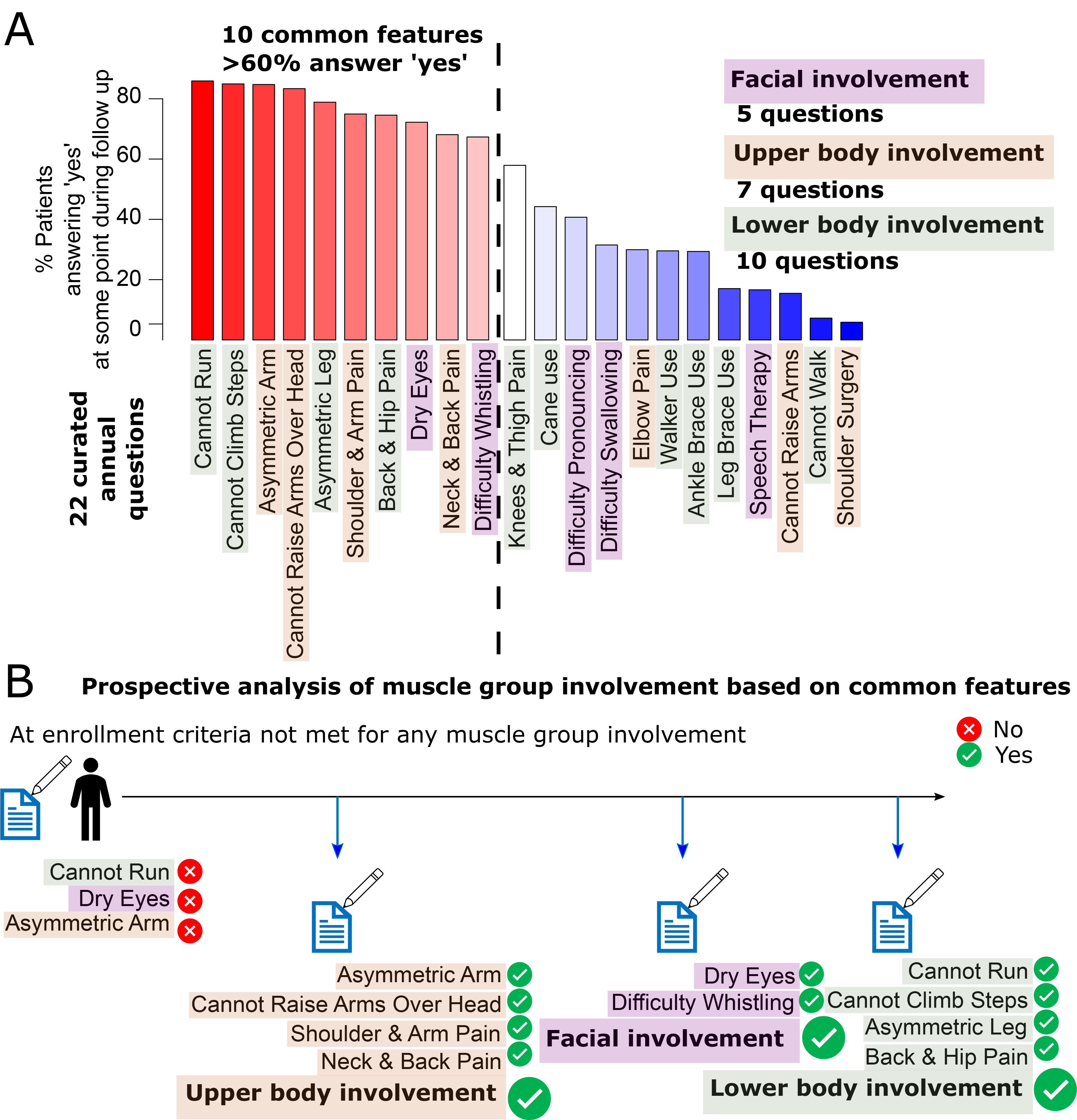
Prevalence of FSHD reported features and approach to outcome assessment. **(A)** Bar chart displays the percentage of the 511 patients with follow up, reporting ‘yes’ to each of the 22 manually curated questions, either at enrolment or during follow up. Questions are outlined in purple boxes to indicate assignment to facial involvement, brown boxes for upper body involvement and grey boxes for lower body involvement. Bars are coloured red if the question is answered ‘yes’ by >60% of patients, and the question is said to describe a common feature, bars are coloured white/blue otherwise. A dotted line separates common features from less common features. **(B)** Schematic outlining how muscle group involvement as an outcome is determined. Patients who submit a questionnaire reporting ‘yes’ to all of the common features associated to a given muscle group are labelled as having involvement of the muscle group at that point.

Given their relative prevalence, we considered these 10 common variables as representative of muscle group involvement for prospective analysis. We labelled a patient as experiencing facial involvement once they have completed a questionnaire reporting ‘yes’ to the two common variables attributed to the face, as experiencing upper body involvement after reporting ‘yes’ to the four common variables attributed to the upper body, and as experiencing lower body involvement following reporting ‘yes’ to the four common variables attributed to the lower body (**Fig 2B**).

Of the 511 FSHD1 patients, 113 (22.1%) never met the criteria for involvement of any muscle group throughout follow up, while 21 (4.1%) enrolled meeting criteria for involvement of all muscle groups (**Table 1**). The remaining 377 (73.7%) FSHD1 patients acquired involvement of at least one muscle group during follow up, allowing us to assign an age of involvement for one or more muscle groups, in the majority of patients.

**Table 1.** Numbers and proportions of patients experiencing involvement of each muscle group. For each muscle group (facial, upper/lower body), the number (alongside percentages in brackets) of patients reporting involvement at enrolment, during follow up and not at all are displayed.

| No lower body involvement |  |  |  |  |
| --- | --- | --- | --- | --- |
|  | No upper body involvement | Upper body involvement during follow up | Upper body involvement at enrollment | Total |
| No facial involvement | 113 (22.1%) | 20 (3.9%) | 21 (4.1%) | 154 (30.1%) |
| Facial involvement during follow up | 23 (4.5%) | 5 (1.0%) | 6 (1.2%) | 34 (6.7%) |
| Facial involvement at enrollment | 28 (5.5%) | 5 (1.0%) | 9 (1.8%) | 42 (8.2%) |
| Total | 164 (32.1%) | 30 (5.9%) | 36 (7.0%) | 230 (45.0%) |

| Lower body involvement during follow up |  |  |  |  |
| --- | --- | --- | --- | --- |
|  | No upper body involvement | Upper body involvement during follow up | Upper body involvement at enrollment | Total |
| No facial involvement | 15 (2.9%) | 21 (4.1%) | 10 (2.0%) | 46 (9.0%) |
| Facial involvement during follow up | 20 (3.9%) | 21 (4.1%) | 10 (2.0%) | 51 (10.0%) |
| Facial involvement at enrollment | 13 (2.5%) | 10 (2.0%) | 10 (2.0%) | 33 (6.5%) |
| Total | 48 (9.4%) | 52 (10.2%) | 30 (5.9%) | 130 (25.4%) |

| Lower body involvement at enrollment |  |  |  |  |
| --- | --- | --- | --- | --- |
|  | No upper body involvement | Upper body involvement during follow up | Upper body involvement at enrollment | Total |
| No facial involvement | 23 (4.5%) | 9 (1.8%) | 11 (2.2%) | 43 (8.4%) |
| Facial involvement during follow up | 14 (2.7%) | 16 (3.1%) | 27 (5.3%) | 57 (11.2%) |
| Facial involvement at enrollment | 16 (3.1%) | 14 (2.7%) | 21 (4.1%) | 51 (10.0%) |
| Total | 53 (10.4%) | 39 (7.6%) | 59 (11.5%) | 151 (29.5%) |

### Cigarette smokers with FSHD have double the risk of facial and lower body involvement

For each of our 3 muscle groups separately, we performed a time-to-event analysis. Employing multivariate Cox proportional hazards models, we modelled age of involvement of each muscle group as a function of the 7 explanatory variables determined at enrolment: D4Z4 repeat length, sex, BMI, self-reported diabetes, hypertension, high cholesterol and cigarette smoking (**Fig 3A**). Patients meeting criteria for muscle group involvement at enrolment were excluded from analysis of that muscle group. Patients who never met the criteria for involvement of a given muscle group were censored at the age of completion of their most recent follow up questionnaire. Patients in each muscle group analysis experiencing each explanatory variable is detailed (**Supplementary Table S2)**.

**Fig. 3.**
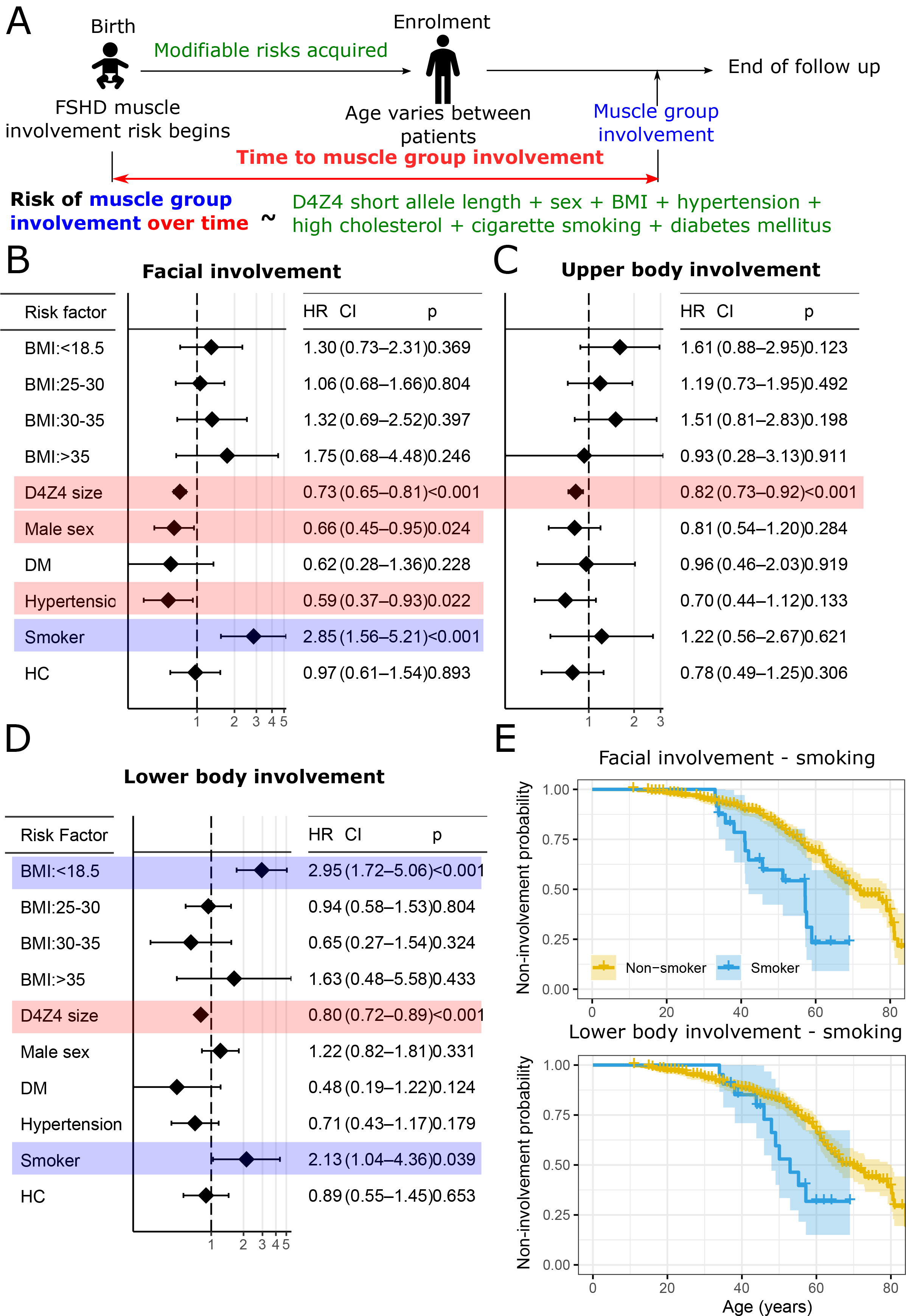
Time-to-event analysis identifies smoking as associated with double the risk of facial and lower limb involvement. **(A)** Schematic outlining time-to-event analysis, each muscle group (facial, upper/lower body) is considered separately, only patients who experience muscle group involvement after enrolment are considered. Patients are considered at risk of involvement from birth, potential risk factors are reported at enrolment and thus prior to involvement. Age of involvement of each muscle group is modelled as a multivariate function of the 7 explanatory variables, via Cox proportional hazards models. Forrest plots display the hazard ratios for each of the 7 explanatory variables as a risk factor for **(B)** facial involvement **(C)** upper body involvement **(D)** lower body involvement. A diamond denotes the point estimate of the hazard ratio and whiskers display the 95% confidence interval, variables for which the whiskers cross the line hazard ratio=1 are not significantly associated with risk of muscle group involvement. Numerical values for the hazard ratio, 95% confidence interval and *p*-value, for each variable are displayed alongside each plot. BMI hazard ratios are reported compared to a normal BMI of 18.5-25. D4Z4 size hazard ratios are reported as the added risk of an additional 3.3kb D4Z4 unit. All other variable hazard ratios are reported as the added risk of reporting ‘yes’ to having that variable. Blue boxes outline variables associated with increased risk of muscle involvement and red boxes decreased risk. HC denotes high cholesterol, DM denotes diabetes mellitus. **(E)** Kaplan Meier plots display the age of facial and lower body involvement for cigarette smokers (blue) compared to non-smokers (yellow). A tick denotes a censored data point and 95% confidence intervals for smokers and non-smokers are outlined.

We found that longer D4Z4 truncated arrays associated with later involvement of all 3 muscle groups (facial, HR per 3.3kb unit: 0.73(0.65,0.81), *p*=2.7x10^-8^, upper body, HR per 3.3kb unit: 0.81(0.73,0.91), *p*=3.9x10^-4^, lower body, HR per 3.3kb unit: 0.80(0.72,0.89), *p*=6.7x10^-5^, **Fig 3B-D**), consistent with the literature (2, 29, 31). Low BMI was also a significant risk factor for earlier lower limb involvement (HR compared to normal BMI: 3.16(1.85,5.38), *p*=2.4x10^-5^), in line with a recent machine learning analysis of the USA FSHD registry that identified low BMI as a risk factor for earlier wheelchair use (26). We also identified female sex as a significant risk factor for earlier facial involvement in our data set (HR: 1.54(1.06, 2.22), *p*=0.02). Curiously, hypertension was associated with later facial involvement (HR: 0.58(0.37, 0.92), *p*=0.02), but was not associated with upper or lower body involvement.

Importantly, we found that cigarette smoking was associated with more than double the risk of facial and lower limb involvement in our FSHD patients (facial, HR: 2.82(1.54,5.17), *p*=7.6x10^-4^ lower body, HR: 2.07(1.01,4.26), *p*=0.047, **Fig 3E**).

### FSHD muscle cells are disproportionally vulnerable to cigarette smoke-induced mitochondrial stress and cellular apoptosis

Cigarette smokers with FSHD1 in our dataset progress faster than non-smokers, and cigarette smoke contains a number of oxidative stressors (40). FSHD muscle cells suffer from oxidative stress sensitivity through mitochondrial dysfunction (15, 19, 41, 42), and are thus theoretically more susceptible to cell death in response to exogenous oxidative stressors (22). We exposed patient-derived FSHD models to CSM to test whether FSHD clones show differential responses compared to matched controls (**Fig 4A**). Exposure to CSM reduced the cell proliferation rate in FSHD and unaffected clones alike, as assessed by EdU incorporation (**Fig 4B, C**) and DNA quantitation (**Fig 4D**). However, FSHD clones (54-12, 16A) showed a significantly greater reduction than unaffected clones (54-6, 16U) on fold change to the respective untreated control. Likewise, FSHD myoblasts also exhibited disproportionally higher apoptosis levels upon long term exposure to CSM compared to unaffected controls (**Fig 4E**).

**Fig. 4.**
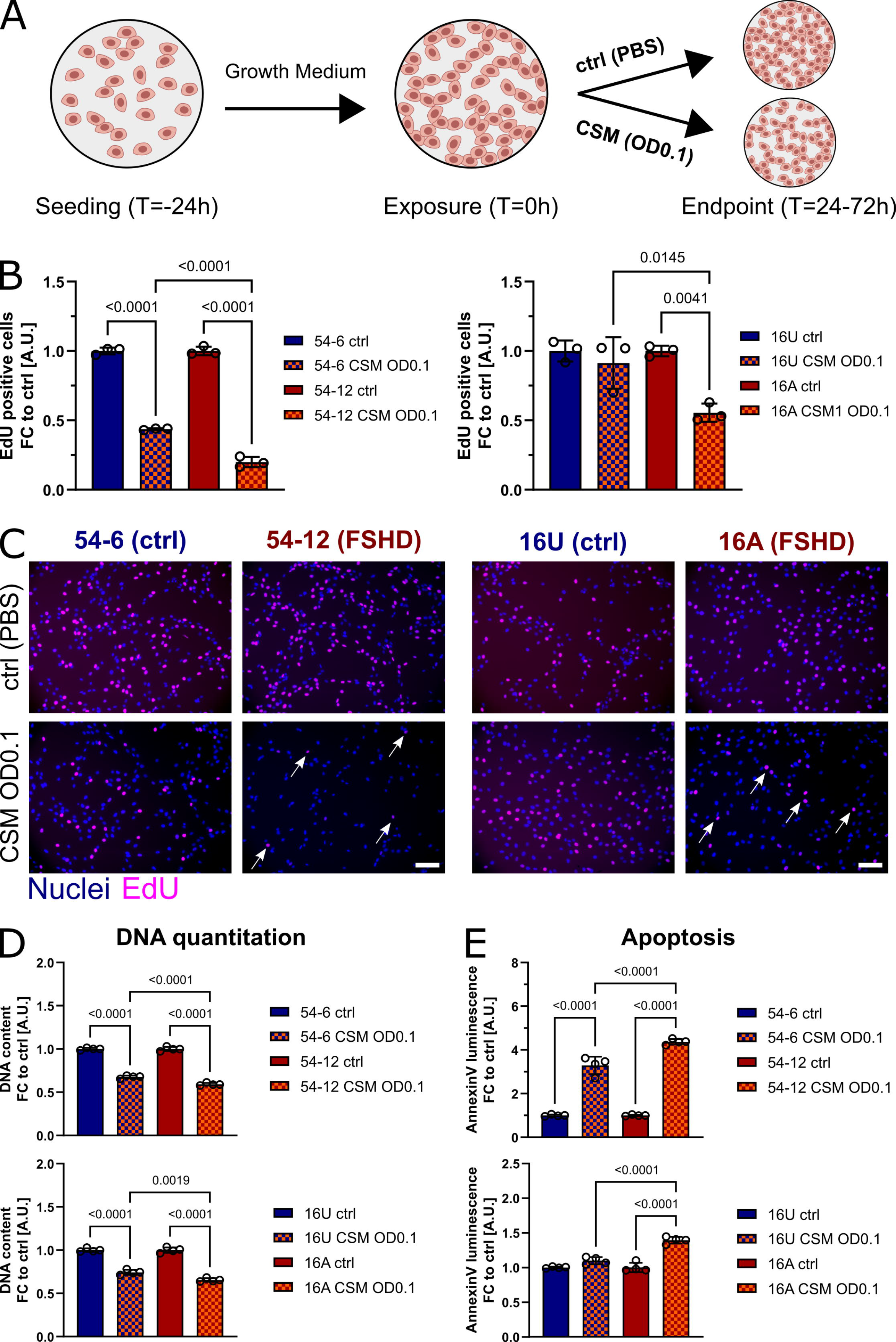
FSHD myoblasts show increased susceptibility to CSM-induced apoptosis. **(A)** Experimental setup: FSHD (54-12, 16A) and matched control myoblasts (54-6, 16U) were exposed to CSM (OD0.1) 24h post-seeding, with subsequent endpoint assessment of proliferation (EdU), cell number (DNA quantitation) and apoptosis after 24h and 72h, respectively. **(B)** Exposure to CSM for 24h reduces proliferation (percentage of EdU-positive nuclei) disproportionally in FSHD myoblasts (red) compared to controls (blue). **(C)** Representative immunofluorescence micrographs after 24h exposure to CSM (nuclei = blue, EdU = purple; scale bar represents 100μm, arrows indicate EdU-positive cells). **(D)** CSM reduces cell number (quantified as DNA content through HOECHST33342 fluorescence after exposure to CSM for 24h) and induces **(E)** apoptosis (AnnexinV luminescence after exposure to CSM for 72h) in FSHD myoblasts (red) to a higher degree than in controls (blue). n=3-4, data is mean ± s.d. with p values as indicated.

Since a variety of compounds in cigarette smoke alter mitochondrial respiratory chain activity by directly inhibiting mitochondrial complexes (43–46), we also investigated changes in mitochondrial ROS (mitoROS) metabolism after CSM exposure. CSM triggered an immediate reduction of mitochondrial metabolic activity, which again was more pronounced in FSHD myoblasts (**Fig 5A**). Interestingly, mitochondria in FSHD myoblasts responded to CSM exposure with disproportionally higher mitoROS levels (**Fig 5B**) and membrane potential (ΔΨm) (**Fig 5C**), the latter a known modulator of mitoROS production from the respiratory chain. This suggests that mitochondria in FSHD cells are more vulnerable than healthy myoblasts to CSM- induced impairment of the respiratory chain, leading to more pronounced oxidative stress through mitoROS.

**Fig. 5:**
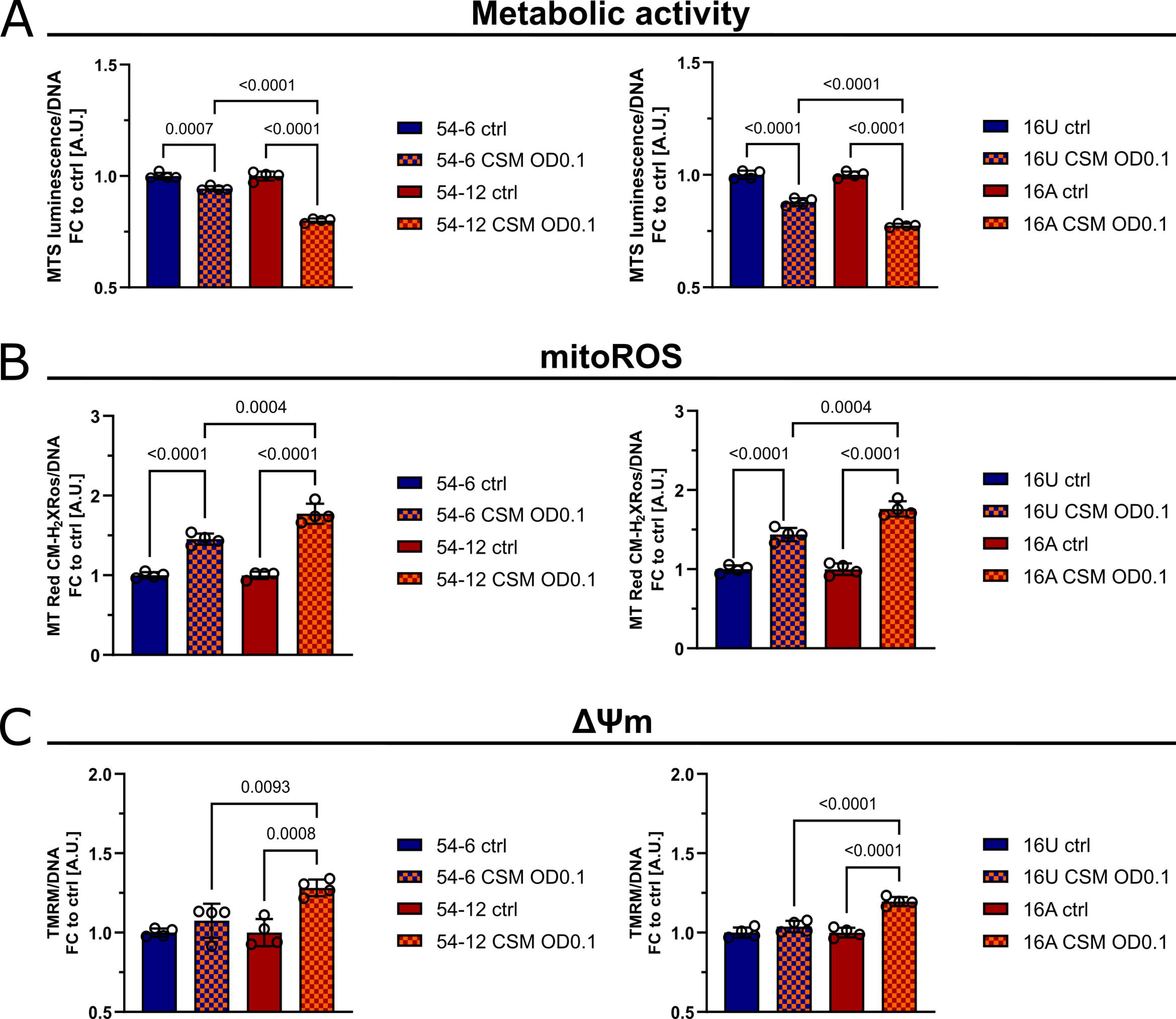
FSHD myoblasts are more susceptible to CSM-induced mitochondrial stress. **(A)** Reduction of mitochondrial metabolic activity is an early event in the response to CSM (6h exposure), with FSHD myoblasts (red) more severely affected than controls (blue). **(B)** Elevated mitoROS levels in FSHD myoblasts (red), driven by more pronounced increase of **(C)** ΔΨm in FSHD mitochondria, compared to controls (blue) after 24h exposure to CSM. n=4, data is mean ± s.d. with p values as indicated.

### CSM exacerbates myotube hypotrophy in FSHD

Redox regulation and mitochondrial function play multifaceted and overlapping roles in skeletal myogenesis and muscle regeneration/homeostasis (47, 48) and we found disproportionate changes in mitoROS metabolism in FSHD myoblasts after exposure to CSM. FSHD myoblasts differentiate into hypotrophic myotubes (18), and alleviation of (mitochondrial) oxidative stress rescues this hypotrophic myotube phenotype (15).

We thus subjected FSHD myoblasts to myogenic differentiation in the presence of CSM at varying concentrations alongside matched controls, to test for differential susceptibility to CSM- induced hypotrophy (**Fig S1A**). Low concentrations of CSM (OD0.01) had no overt morphological effect on myogenesis, whereas medium (OD0.05) and high (OD0.1) concentrations visibly affected myogenesis, with the high almost completely blocking myotube formation (**Fig S1B**). Therefore, we chose a medium concentration of CSM (OD0.05) for myotube analysis. Exposure to CSM over the course of differentiation (72h, **Fig 6A**) impaired myogenesis to a greater extent in FSHD myoblasts (54-12, 16A) than in matched controls (54-6, 16U; **Fig 6B**): both MyHC-positive myotube area, used as a readout for myotube hypotrophy (**Fig 6C**), and myogenic differentiation index (**Fig 6D**) were disproportionally reduced in FSHD myotubes. This suggests that CSM-induced metabolic stress aggravates FSHD pathology in myotubes.

**Fig. 6:**
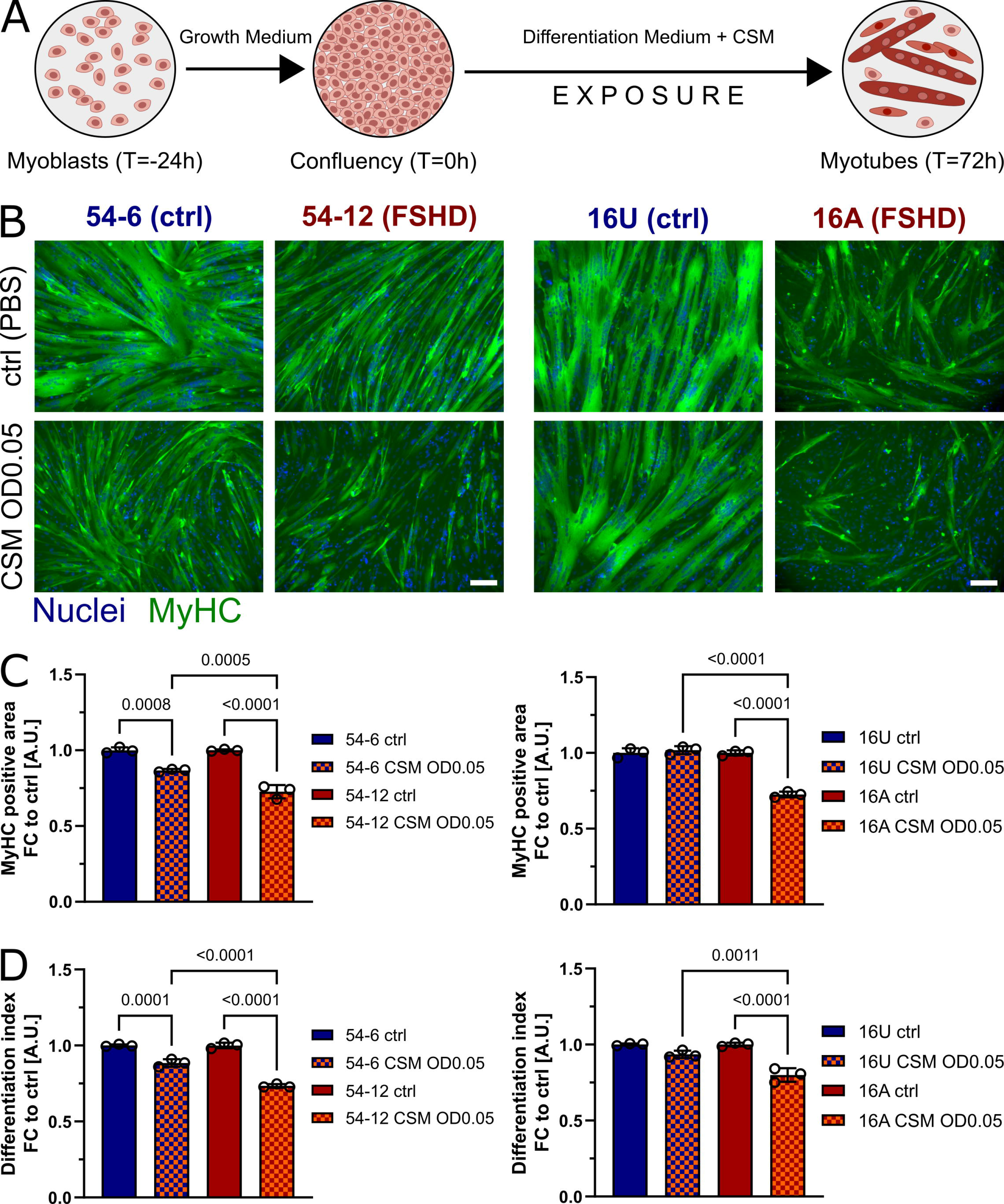
FSHD myogenesis is sensitive to CSM-induced myotube hypotrophy. **(A)** Experimental setup: FSHD (54-12, 16A) and matched control myoblasts (54-6, 16U) were grown to confluency, and induced to differentiate into myotubes under control conditions (PBS) or exposure to CSM (OD0.05) for 72h. **(B)** FSHD myotubes (red) produce a hypotrophic myotube phenotype compared to controls (blue), which is disproportionally aggravated by exposure to CSM in FSHD myotubes (nuclei = blue, MyHC = green, scale bar represents 100μm). **(C, D)** Quantitation of MyHC-positive area (as readout for myotube hypotrophy) and differentiation index (percentage of myonuclei within the MyHC-positive area) from representative immunofluorescence micrographs after differentiation in the presence of CSM compared to untreated (PBS) controls. n=3, data is mean ± s.d. with p values as indicated.

Together, our data suggests that FSHD muscle cells are more susceptible to CSM-induced metabolic disturbances, which aggravates FSHD pathologic phenotypes in myoblasts and myotubes.

## DISCUSSION

We performed a prospective analysis of the complex, multimodal, longitudinal USA FSHD registry dataset, comprising 511 FSHD1 patients assessed annually for an average of 8 years. By manual curation of 189 baseline features and 37 annually assessed FSHD severity features, we developed an FSHD-informed approach to assessing the impact of multiple modifiable factors on the risk of developing facial, upper and lower body involvement in FSHD.

Our prospective cohort study revealed that cigarette smoking associated with double the risk of facial and lower body involvement in FSHD over time, independently of other factors.

This corresponds to cigarette smokers experiencing important functional deficits including inability to run and climb steps without using a handrail, pain in the hip and lower back and asymmetric leg weakness at significantly younger ages than non-smoking patients with FSHD1.

Cigarette smoking is an important modifiable risk factor for a wide spectrum of pathology including lung cancer, chronic obstructive pulmonary disease and coronary heart disease (49, 50). Smoking cessation approaches range from risk counselling to nicotine replacement therapy, and represent an important part of clinical management for these conditions. Our findings suggest that smoking cessation should also be promoted in clinical management of FSHD, and provide evidence to aid risk counselling.

Vincenten et al., 2021, performed a cross-sectional study of 198 Dutch FSHD patients and found no association between smoking and disease severity (39). In our prospective cohort study of 511 USA FSHD patients, we ascertained smoking status prior to development of muscle group involvement and monitored how risk of such involvement differed between FSHD1 smokers and non-smokers (adjusting for other factors). This approach is more sensitive in identification of smoking as a risk factor, compared to time-independent multivariate regression employed in a cross-sectional study. Moreover, Vincenten et al. assessed FSHD clinical severity via the Lamperti score (51). The Lamperti score assigns points from 0-15 based on a combined clinical assessment of multiple muscle groups in an FSHD patient, 2 points are reserved for facial involvement, 5 for upper body, 7 for lower body and 1 for abdominal. We curated temporal information on patient function, pain, weakness distribution and use of external aids to gain a dynamic understanding of facial, upper body and lower body involvement separately in FSHD. We found that risks of both facial and lower body involvement were enhanced in FSHD1 cigarette smokers, while the risk of upper body involvement was unaltered. This motivates a more muscle group targeted approach to FSHD severity assessment.

The USA FSHD registry employed a mix of chart review and an annual questionnaire populated with self-reported patient features. Our analysis clearly reproduces well known FSHD features, such as shorter D4Z4 arrays associating with greater risk of progression but self-reported data may be subject to bias. A recent analysis of the French FSHD registry directly investigated concordance between self-reported and clinician-reported FSHD features (52). There was a strong concordance between patients and clinicians completing questionnaires assessing lower body (Vignos score) and upper body (Brooke score) function, as well as muscle pain (Visual Analogue Pain Score). However, facial involvement (eye closure and whistling ability) was recorded significantly more by clinicians than patients. Self-reported assessments of upper and lower body involvement we employ are likely representative of true clinical assessment, while facial involvement may be an underestimate. Facial muscle dysfunction may also be more readily noticed by smokers due to the more frequent use during smoking. It is important therefore, that we find cigarette smoking associated not only with increased risk of facial involvement in FSHD but also lower limb involvement. A limitation of the USA FSHD registry, is that pack years smoked was not recorded. Further data thus needs to be collected to establish a dose dependent link between cigarette smoking and FSHD disease progression.

Our cell data suggest that vulnerability to cigarette smoke is increased in FSHD patient-derived muscle cells. A single acute exposure to CSM triggered a disproportionate reduction in cell viability and proliferation as well as a more pronounced myogenic differentiation deficit in FSHD muscle cells *in vitro,* compared to unaffected controls - both in isogenic and sibling-matched models. Interestingly, CSM-induced mitochondrial dysfunction, evident as decreased mitochondrial metabolic activity with concomitant increase in mitoROS levels, was more pronounced in FSHD myoblasts, and driven by a (likely transient) increase of mitochondrial membrane potential, which is known to eventually collapse as mitochondrial dysfunction and bioenergetic crisis progress (53). This indicates that FSHD muscles may have lower intrinsic tolerance to cigarette smoking-induced metabolic/oxidative stress and atrophy.

Complexity of cigarette smoke composition has been extensively studied over decades, with more than 8400 chemical constituents identified (54). Mitochondria are very sensitive to environmental toxicants, with haeme groups in the respiratory complexes particularly prone to inactivation/inhibition by oxidants and/or carbon monoxide. Cigarette smoke contains many redox active compounds and thus profoundly affects mitochondrial metabolism through changes in respiratory chain activity, mitoROS generation, mitochondrial DNA (mtDNA) stability, mitophagy flux and (mitochondrial) redox signalling (reviewed by: (55)). Cigarette smoke is known to inhibit complex IV (cytochrome C oxidase) (43, 56), and the large part of this effect has been attributed to cyanide (57) **and carbon monoxide** (58). Therefore, cigarette smoking likely accelerates FSHD pathology in multiple ways. After the seminal observation that myoblasts from FSHD patients are more susceptible to oxidative stress (22), we and others have identified dysfunctional mitochondria as a major source of oxidative stress/damage in FSHD muscle cells (15, 41, 42). Decreased complex IV activity has been observed in muscle fibres from FSHD patient biopsies (19), and we recently reported that DUX4 overexpression elicits mitochondrial dysfunction in a complex I-dependent manner (15). Nicotine specifically inhibits complex I-linked respiration in isolated murine skeletal muscle mitochondria (59), therefore mitochondrial dysfunction and subsequent oxidative damage in FSHD muscle is likely further aggravated through cigarette smoke constituents acting on complex I and IV.

DUX4 underlies transcriptional redox regulation (60) and regulates around 200 of its target genes indirectly through ROS (41). DUX4 expression in FSHD-derived iPSCs is also enhanced by oxidative stress (60). Thus, cigarette smoke constituents may directly or indirectly enhance DUX4 toxicity through elevated mitoROS generation and subsequent modulation of DUX4 transcription, protein stability and target gene activation.

Apart from mitochondrial respiratory chain inhibition, smoking also increases blood carboxyhaemoglobin levels which reduces oxygen delivery to muscles (61). This is usually accompanied by increased oxidative stress through Fenton reactions and triggers ferroptosis (62).

Impaired muscle oxygen demand during exercise in FSHD patients (63) could be a consequence of disturbed iron homeostasis. In addition, perturbance of the molecular and metabolic response to hypoxia in FSHD muscle cells has been identified (15, 64), with a direct pathomechanistic link between DUX4 toxicity and hypoxia-inducible factor 1α (HIF1α) (65).

Since mitochondria are the main site of oxygen consumption, haeme metabolism and ROS generation, it is likely that disproportionate impairment of oxidative metabolism is a main route by which cigarette smoking affects FSHD patients, even though precise mechanisms of mitochondrial dysfunction, such as potential reverse electron transfer and/or ATP hydrolysis via complex V reversal, are still insufficiently understood.

Cigarette smoking also induces muscle weakness via p38 activation of NF-κB, driving expression of atrophy inducing MuRF-1 (40). Concordantly, MuRF-1 is elevated in FSHD patient myotubes and DUX4 over-expressing myotubes (66, 67), while NF-κB activation associates with FSHD oxidative stress sensitivity (68), implying a molecular sensitivity to smoking induced muscle weakness in FSHD. Moreover, as a p38α inhibitor is in clinical trials for FSHD (69), cigarette smoking could directly counteract positive benefits on FSHD muscle. Cigarette smoking also activates pro-inflammatory cytokines TNFα and IL-6 (40), both of which are elevated in FSHD patient serum and inversely correlate with muscle strength (19, 64). Lastly, the degree of D4Z4 hypomethylation associates with FSHD severity (5, 70), and smoking is linked to genome wide de-methylation which could further contribute (71).

In summary, we report cigarette smoking is associated with a two-fold increased risk of facial and lower limb involvement in a prospective cohort of 511 USA FSHD1 patients. We further demonstrate that cigarette smoke medium disproportionately impairs proliferation, viability and myogenic differentiation of FSHD patient-derived myoblasts, in a manner directly linked to mitochondrial dysfunction. Our findings provide the first evidence to support smoking cessation as part of the clinical management plan to reduce the rate of disease progression in FSHD.

## MATERIALS AND METHODS

### The USA FSHD Registry

The USA FSHD data has been previously described (26, 72). Briefly de-identified data was collected on individuals participating in USA FSHD Registry located at the University of Rochester Medical Center in Rochester, New York (https://www.urmc.rochester.edu/neurology/national-registry.aspx). Data analysed were collected from January 2002 to September 2019. All patients considered in this study were genetically confirmed FSHD1.

578 FSHD1 patients were enrolled in the registry, of which 511 had at least one annual follow up questionnaire, with a mean follow up of 8 years, range 1-17 years. At enrolment, 189 variables were assessed covering a wide range of demographic and FSHD-related outcomes. Including, but not limited to D4Z4 small allele size, sex, BMI, self-reported hypertension, diabetes mellitus, high cholesterol and cigarette smoking status. At each annual follow-up a questionnaire was completed including 37 yes/no questions describing pathological involvement of various muscle groups.

### Manual curation of annual follow up questionnaire

37 yes/no questions answered in the annual questionnaires were manually curated to assign an unambiguous muscle group and ensure that an answer of ‘yes’ implied disease progression.

15 questions were excluded, due to ambiguity. 9 questions referred to mobility and transfers, describing function of multiple muscles and could not be assigned unambiguously to a single muscle group. Similarly, wheelchair use can be driven by involvement of muscle multiple groups, and so was also excluded. 5 questions assigned to upper and lower body involvement described intermediate progression of inability raise arms and ambulate respectively, for these questions a ‘no’ could imply either an earlier stage of disease or a later stage, these questions were thus excluded.

Assignment of the remaining 22 questions was as follows:

Facial involvement (5 questions):

Are your eyes dry/irritated?

Do you have trouble whistling or drinking through a straw? Do you have difficulty pronouncing certain words?

Do you have difficulty swallowing? Have you ever received speech therapy?

Upper body involvement (7 questions):

Are you unable to raise your arms up sideways over your head? Are you unable to raise your arms sideways at all?

Is one arm noticeably more affected by the disease?

Do you have muscle or joint pain in (a) neck/upper back? (b) shoulder/upper arms? (c) elbows? Have you had surgery to fix your shoulder blades?

Lower body involvement (10 questions):

Are you unable to run?

Are you unable to climb stairs without using handrail or cane? Are you unable to walk?

Is one leg noticeably more affected by the disease?

Do you have muscle or joint pain in (a) lower back/hips (b) knees/thighs?

Do you require the use of (a) ankle braces? (b) long leg braces? (c) a cane (d) a walker

The proportion of the 511 patients with follow up, who answered ‘yes’ to each of these questions either at enrolment or at annual follow up was determined, questions answered by more than 60% of participants were considered common events.

### Time-to-event analysis

Time-to-event analysis was performed using the *survivalAnalysis* package in R (73).

Three outcomes were analysed separately: onset of facial, upper body or lower body muscle involvement.

Onset of facial involvement was assessed as the age of receipt of the first questionnaire reporting ‘yes’ to 2 common questions: ‘Are your eyes dry/irritated?’ and ‘Do you have trouble whistling or drinking through a straw?’

Onset of upper body involvement was assessed as the age of receipt of the first questionnaire reporting ‘yes’ to 4 common questions: ‘Are you unable to raise your arms up sideways over your head?’ and ‘Is one arm noticeably more affected by the disease?’ and ‘Do you have muscle or joint pain in neck/upper back?’ and ‘shoulder/upper arms?’.

Onset of lower body involvement was assessed as the age of receipt of the first questionnaire reporting ‘yes’ to 4 common questions: ‘Are you unable to run?’ and ‘Are you unable to climb stairs without using handrail or cane?’ and ‘Is one leg noticeably more affected by the disease?’ and ‘Do you have muscle or joint pain in lower back/hips?’

For each muscle group separately, if involvement occurred at/before joining the registry the patient was excluded, if involvement did not occur during follow up the patient was censored at the age of most recent questionnaire. A Cox proportional hazards model was fit modelling the age of involvement of each muscle group as a function of 7 variables assessed at enrolment (and thus recorded before involvement): D4Z4 repeat length, sex, BMI, self-reported diabetes, hypertension, high cholesterol and smoking.

Wald and Logrank tests determined overall model significance and *z*-scores describing hazard ratios determined independent associations of covariates. Significance was assessed at the 5% level.

### Multivariate regression

Multivariate linear regression determined independent associations between 7 explanatory variables: D4Z4 repeat length, sex, BMI, self-reported diabetes, hypertension, high cholesterol and smoking. Each variable was modelled as a linear combination of the remaining 6, resulting in 21 unique coefficients and *p*-values. Adjustment for multiple testing of 21 *p*-values was performed via Benjamani-Hochberg method. Significance of adjusted *p*-values was assessed at the 5% level. These results are reported in **Supplementary Table S1**.

### Cell Culture

The FSHD patient-derived cellular models used in this study were: the isogenic ‘54’ series derived from the biceps of a male mosaic FSHD1 patient (74), where 54-6 (13 D4Z4 repeats) is the uncontracted control clone and 54-12 (3 D4Z4 repeats) the contracted FSHD clone; and the ‘16’ series, a sibling-matched immortalised model derived from biceps muscle (75) where 16A is the D4Z4 contracted FSHD line and 16U the uncontracted control line from a first-degree relative.

Human myoblast lines were maintained in Skeletal Muscle Cell Growth Medium (Promocell, Heidelberg, Germany) supplemented with 20% foetal bovine serum (FBS; ThermoFisher Scientific, MA, USA), 50 μg/ml fetuin (bovine), 10 ng/ml epidermal growth factor (recombinant human), 1 ng/ml basic fibroblast growth factor (recombinant human), 10 μg/ml insulin (recombinant human), 0.4 μg/ml dexamethasone (all supplemented as PromoCell SupplementMix) and 50 μg/ml gentamycin (Sigma Aldrich) in a humidified incubator at 37°C with 5% CO2. Differentiation was induced by switching confluent myoblast cultures to Dulbecco’s Modified Eagle Medium (DMEM) GlutaMax (ThermoFisher Scientific) supplemented with 0.5% FBS, 10 μg/mL recombinant human insulin (Sigma-Aldrich) and 50 μg/mL gentamycin.

### Preparation of cigarette smoke media (CSM)

1R6F Research grade cigarettes (University of Kentucky) were connected to a 60 ml syringe with rubber tubing. After the cigarette was lit, it was withdrawn through the full length of the syringe, each full draw took an average 9 seconds. Withdrawn smoke was bubbled through 10 ml PBS; every “exhalation” lasted an average of 25 seconds. The process was repeated until the cigarette was fully consumed; an average of 9 draws. A full spectrum analysis was performed before the absorbance was measured. Maximum OD was recorded, and subsequent dilutions were made in PBS. The final working dilution was 1:10 into either high serum medium or low serum medium depending on requirements.

### Assessment of Cell proliferation and DNA quantitation

To assess proliferation via EdU incorporation, 10000 cells/well were seeded into black, clear-bottom polystyrene 96-well plates (Corning®; Sigma Aldrich, St. Louis, US). The next day, CSM was added and cells assayed 24 hours later. EdU incorporation after a 2 hours pulse at the end of the incubation period was assayed using the Click-iT EdU Imaging Kit (Life Technologies, Carlsbad, US) as per manufacturer’s instructions. For DNA quantitation, nuclei were stained with HOECHST33342 (0.5 μg/mL; ThermoFisher Scientific, Waltham, US) for 20 min, and fluorescence intensity measured on a CLARIOStar microplate reader (BMG Labtech, Ortenberg, Germany) in spectral averaging scan mode (100 flashes per well, scan diameter 6 mm).

### Apoptosis assaying

Apoptosis was measured with the non-invasive luminescence RealTime-Glo^TM^ Annexin V Apoptosis assay (Promega, Southampton, UK), according to the manufacturer’s instructions. Briefly, 5000 cells/well were seeded into opaque walled polystyrene 96-well plates (Corning®, Sigma Aldrich). 24 hours later, cells were exposed to CSM and assay substrates were added simultaneously for longitudinal assaying of apoptosis for up to 72 hours. Apoptosis was quantified as luminescence signal intensity, as assessed on a Mithras LB940 multimode microplate reader (0.2s exposure; Berthold Technologies, Bad Wildbad, Germany).

### Metabolic activity analysis

Metabolic activity was assessed with the luminescence RealTime-Glo^TM^ MT Cell Viability assay (Promega) with a modified protocol. 10000 cells/well were seeded into opaque walled polystyrene 96-well plates (Corning®, Sigma Aldrich). 24 hours later, were exposed to CSM and assay substrates were added simultaneously for longitudinal assaying for up to 6 hours.

Luminescence was read with a Mithras LB940 multimode microplate reader (0.1s exposure), and metabolic activity was calculated as the ratio between luminescent signal and DNA content, as assessed through HOECHST33342 (0.5 ug/mL; ThermoFisher Scientific) fluorescence from sister cultures undergoing the same treatment.

### mitoROS and ΔΨm measurements

mitoROS and ΔΨm were measured as recently described (15). Briefly, 25000 cells/well were seeded into black, clear-bottom polystyrene 96-well plates (Corning®; Sigma Aldrich). The next day, CSM was added and mitoROS/ΔΨm assayed 24 hours later. MitoTracker® Red CM- H2XROS (ThermoFisher, Scientific) was used for mitoROS detection (final concentration 5 μM), ΔΨm was assessed by Tetramethylrhodamine methyl ester (TMRM; Sigma Aldrich) staining (final concentration 100 nM). All measurements were performed in Hank’s Balanced Salt Solution (HBSS) supplemented with Ca^2+^ and Mg^2+^ (Sigma Aldrich). Cells were washed twice with HBSS, followed by incubation with the respective fluorescent mitoROS/ΔΨm probe and HOECHST33342 (0.5 μg/mL; ThermoFisher Scientific) in HBSS for 30 min in the dark.

After staining, cells were washed twice with HBSS and fluorescence intensity was measured on a CLARIOStar Omega microplate reader (BMG Labtech) in spectral averaging scan mode (100 flashes per well, scan diameter 6 mm). For normalization to cell number, fluorescence was normalized to DNA content as simultaneously assessed via HOECHST 33342 (0.5 ug/mL; ThermoFisher Scientific) fluorescence in the same well.

### Immunolabelling of cells

Cells were fixed with 4% paraformaldehyde/phosphate-buffered saline (PBS; Alfa Aesar, Heysham, UK) for 15 min, and permeabilised with 0.1% Triton-X/PBS (Sigma Aldrich) for 15 min. Blocking was performed in 5% normal goat serum (GS)/PBS (Dako, Glostrup, Denmark) for 1h. Cells were incubated with primary antibody in 1% GS/PBS on a rocker overnight at 4°C, washed and subsequently incubated with secondary antibody in 1% GS/PBS for 1 hour in the dark at room temperature. Primary antibody was mouse anti-MyHC (1:400; MF-20; Developmental Studies Hybridoma Bank, IA, USA). Secondary antibody was goat anti-mouse IgG (H+L) AlexaFluor-488 (1:500, A-11001, ThermoFisher Scientific). Nuclear counterstain was performed with 0.5 μg/mL HOECHST33342 (0.5 ug/mL; ThermoFisher Scientific) in PBS for 10 min. Immunofluorescence micrographs were acquired on a Zeiss Axiovert 200M epifluorescence microscope using a Zeiss AxioCam HRm and AxioVision 4.4 software (Zeiss, Jena, Germany).

### Quantitation of MyHC-positive area and differentiation index

Image analysis of MyHC immunofluorescence micrographs was performed with a custom-made high-throughput image analysis software, as recently described (18). For quantitation of the MyHC-positive myotube area, at least three fluorescence micrographs were taken per well and subsequently used for analysis. The mean MyHC-positive area per well was calculated as the average from a minimum three representative images per well. Likewise, differentiation index was automatically determined from the same images as the percentage of nuclei within the MyHC-positive myotube area.

### Statistics

Statistical analysis of cell culture results was performed using GraphPad Prism version 9.5.0 for Windows (GraphPad Software, San Diego, CA, USA; www.graphpad.com). Experiments were performed at least 3 independent times, with detailed n numbers given in each figure. Variance between groups was compared using a Brown Forsythe test and revealed no significant difference. Comparison between experimental groups was performed using a one-way analysis of variance (ANOVA) followed by Tukey’s post-test. *p* < 0.05 was considered significantly different, with *p* values indicated for each significant comparison in the figures.

Statistical analysis of the USA FSHD registry was as detailed above.

### Study Approval

Enrolment in the FSHD registry was voluntary, and all participants provided informed consent for de-identified data to be used in future research projects on FSHD at enrolment.

### Data Availability

All de-identified data used in this study are available upon request from the USA FSHD Registry within the remit of informed consent (https://www.urmc.rochester.edu/neurology/national-registry.aspx). All anonymised data, code, and materials used in this study will be made available on reasonable request subject to suitable materials transfer agreements (MTAs).

## Author contributions

Conceptualization: CRSB, PH, PSZ, JS, RT Methodology: CRSB, PH, JS, NK, JH, HBH, CC, MDK

Investigation: CRSB, PH, MDK Visualization: CRSB, PH

Funding acquisition: CRSB, JS, PH, PSZ Project administration: JS, PSZ, CRSB Supervision: JS, RT, PSZ

Writing – original draft: CRSB, PH

Writing – review & editing: CRSB, PH, JS, PSZ

## Acknowledgments

We thank the patients enrolled in the USA FSHD registry for their kind involvement in this study. We would also like to thank: Vincent Mouly and Kamel Mamchaoui of the platform of immortalisation of human cells from the Institut de Myologie and Charles P. Emerson of the Wellstone Muscular Dystrophy Program, University of Massachusetts Medical School, MA, USA, for providing immortalised myoblast lines.

## List of Supplementary Materials

**Supplementary Table S1: Correlations between the 7 explanatory variables.** Table displays results of multivariate regression analysis of interdependencies of our 7 explanatory variables, D4Z4 short array length, sex, BMI, self-reported hypertension, high cholesterol, diabetes mellitus and cigarette smoking, including *t-*values, *p*-values and adjusted *p*-values. Significant associations are highlighted with red *t*-values and yellow adjusted *p*-values. Common associations such as elevated BMI and hypertension are observed.

**Supplementary Table S2: Breakdown of number of patients experiencing muscle group involvement either at enrolment or during follow up by explanatory variable.** Table displays for each of the 7 explanatory variables (sex, D4Z4 short array length, BMI, self-reported hypertension, high cholesterol, diabetes mellitus and cigarette smoking) the number and percentage of patients experiencing facial, upper and lower body involvement, either at enrolment or during follow up, broken down by variable value.

**Supplementary Figure S1: CSM impairs myogenic differentiation in a dose-dependent manner, independent of FSHD background. (A)** Experimental setup: FSHD (54-12, 16A) and matched control myoblasts (54-6, 16U) were grown to confluency, and induced to differentiate into myotubes under control conditions (PBS) or exposure to varying concentrations of CSM (OD0.01, OD0.05, OD0.1) for 72h. **(B)** A low dose of CSM (OD0.01) at the onset of differentiation does not overtly impair myogenic differentiation, whereas higher doses (OD0.05 and OD0.1, respectively) yield a hypotrophic myotube phenotype (nuclei = blue, MyHC = green, scale bar represents 100μm). n=3, with representative immunofluorescence micrographs shown for each experimental condition.

