## Supplementary figures and images for "Symptom onset and cellular pathology in facioscapulohumeral muscular dystrophy is accelerated by cigarette smoking"

### Figure S1

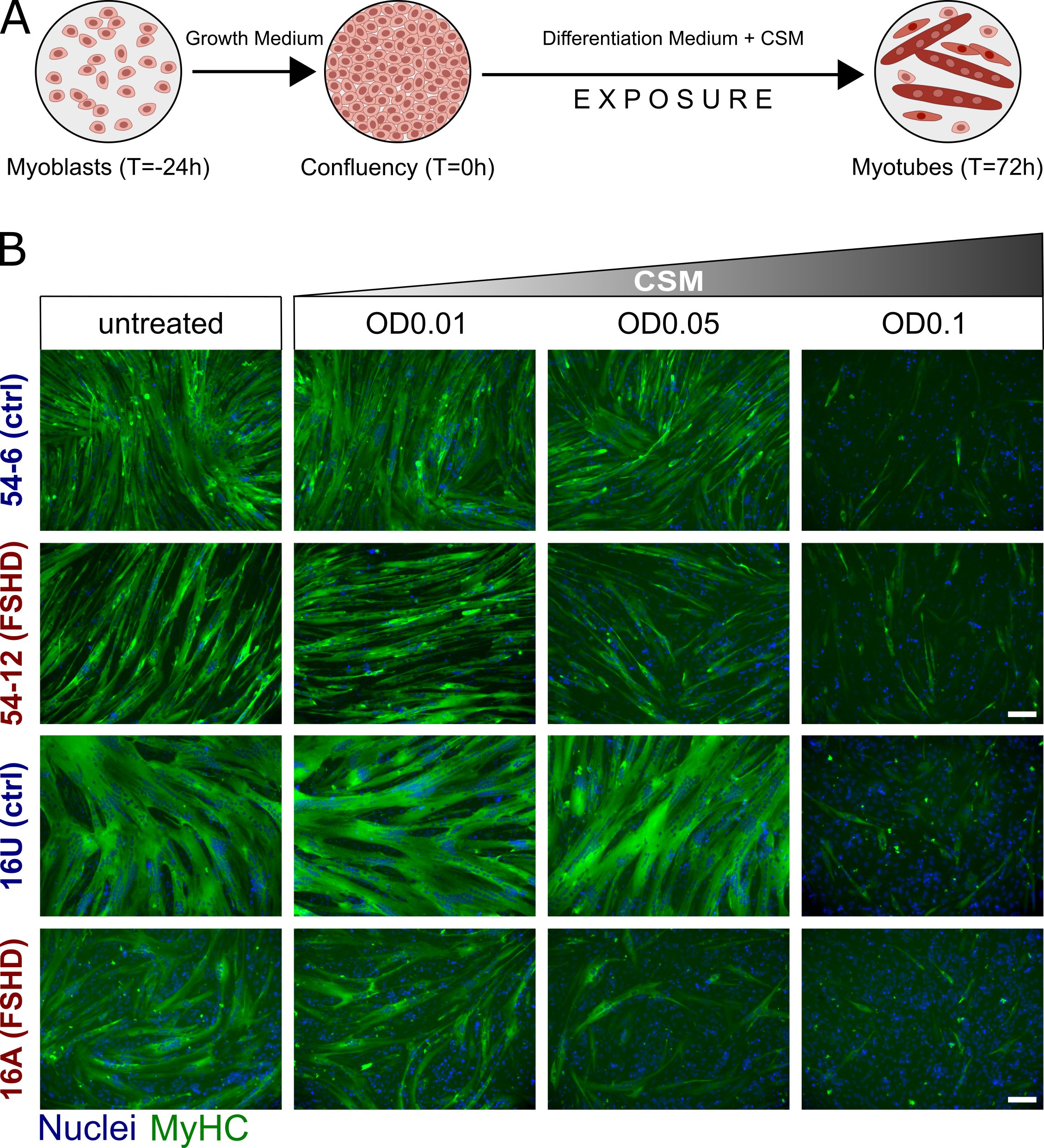
